# Hypertension Phenotypes in a National Database: A Three-Axis State Model Integrating Diagnosis, Treatment Intensity, and Blood Pressure Control (The NDB-K7Ps-Study-8)

**DOI:** 10.64898/2026.07.16.26358276

**Authors:** Kei Nakajima, Airi Sekine

## Abstract

Hypertension is commonly defined as a binary condition despite substantial heterogeneity in diagnosis, treatment, and blood pressure (BP) control. We propose a three-axis state model integrating diagnosis status, treatment intensity, and BP control to better characterize hypertension phenotypes. The framework generates 27 possible states that can be condensed into seven clinically meaningful groups. We applied the model to 5,129,584 Japanese adults using the National Database of Health Insurance Claims and Specific Health Checkups. Hierarchical cluster analysis, sensitivity analysis excluding patients with cardiovascular diseases other than hypertension, and validation against antihypertensive medication use were performed. Overall, 64% of participants were classified as normotensive, whereas 36% belonged to hypertension-related groups, including 11% with unrecognized hypertension and 7% with diagnosed but untreated hypertension. Agreement with data-driven hierarchical cluster analysis was substantial (weighted κ=0.87). The group distribution remained largely unchanged in the sensitivity analysis, supporting the robustness of the proposed classification. Hypertension diagnosis also showed high validity, with a sensitivity of 96.5%, specificity of 91.8%, and substantial agreement with antihypertensive medication use (κ=0.78). This three-axis framework provides a robust and clinically interpretable approach for characterizing hypertension phenotypes, enabling systematic identification of care gaps and supporting research, clinical decision-making, and population health management.

## INTRODUCTION

Hypertension remains one of the leading modifiable risk factors for cardiovascular disease, including myocardial infarction and stroke. Despite decades of research and well-established guidelines, blood pressure (BP) control at the population level remains suboptimal [1]. A key limitation in current epidemiological and clinical frameworks is the reliance on binary definitions of hypertension, whereby individuals are simply classified as normotensive or hypertensive.

Such binary approaches obscure the heterogeneity that exists in real-world care. In clinical practice and population-based data, there is often discordance between diagnostic labeling, pharmacological treatment, and measured BP levels [2]. Several classifications are necessary to reflect diversity in the real world. Individuals with elevated BP may remain undiagnosed, those who are diagnosed may not be treated, and those receiving treatment may continue to have uncontrolled hypertension. Conversely, some individuals receive antihypertensive medication without a recorded diagnosis because of coding variability or alternative indications [3,4]. These discrepancies are meaningful features of healthcare delivery and not merely measurement errors. We suggest that these deviations should be explicitly modeled rather than ignored. Various problems arise when investigating hypertension as an outcome, particularly in research using healthcare data and surveys of the prevalence of hypertension based on a binary definition. However, determining the phenotypic classification that accurately reflects the actual state of hypertension is not easy and requires the efforts of many specialized medical staff. Furthermore, it is necessary to analyze groups with a similar age range, social environment, and ethnicity using the same population-data and diagnostic criteria, rather than simply piecing together the results of multiple studies. Therefore, we propose the following three-axis state model for use in research on hypertension, particularly when using healthcare data, based on a specific algorithm.

### THREE-AXIS STATE MODEL

#### Diagnosis of hypertension and its severity

0: No diagnosis of hypertension

1: Hypertension or primary/essential hypertension (ICD-10; I10).

2: Hypertension with complications (ICD-10; H350, D350, F453, H208, I110, I119, I120, I129, I139, I619, I674)

Secondary hypertension (ICD-10; D350, I150, I151, I152, I159, I270, I272) is excluded from this proposed system to reduce complexity.

#### Treatment intensity

0: No antihypertensive medication

1: Monotherapy (one antihypertensive agent)

2: Combination therapy (two or more antihypertensive agents)

Antihypertensive medication includes angiotensin-converting enzyme inhibitors, angiotensin II receptor blockers, calcium channel blockers, diuretics, β-blockers, α-blockers, and centrally acting agents. At present, 1,974 kinds of these agents are available and marketed as patented, generic, or combination products.

#### Blood pressure level

0: Normal

1: Mild elevation (systolic BP 140–159 mmHg and/or diastolic BP 90–99 mmHg)

2: Severe elevation (systolic BP ≥160 mmHg and/or diastolic BP ≥100 mmHg)

BPs were measured objectively as office BP using an automated BP monitoring device at a medical facility. The number of times BP was measured is unknown, although multiple measurements are recommended in Japan [5].

### CLINICALLY MEANINGFUL GROUPS

This three-dimensional structure yields 27 distinct states (3×3×3), each with an identifier known as the DTB (D, diagnosis status; T, treatment intensity; B, BP level). These 27 states (DTB 000, 001, 002, 010, 011, 012, 020, 021, 022, 100, 101, 102, 110, 111, 112, 120, 121, 122, 200, 201, 202, 210, 211, 212, 220, 221, 222) represent all possible combinations of disease labeling, treatment, and physiological status. However, while this 27-state structure (classification) provides granular detail, it may be impractical for routine interpretation.

Therefore, we propose collapsing these states into seven clinically meaningful tentative groups as follows.

**Group 1 (G1):** A normotensive reference group without diagnosis, treatment, or elevated BP (DTB 000).

**Group 2 (G2)**: Unrecognized hypertension, including two subgroups.

**G2-1** Individuals with elevated BP but no formal diagnosis. Although home BP readings are unknown, white coat hypertension is likely. This subgroup may represent missed opportunities for detection and early intervention [6] (DTB 001, 002).

**G2-2** Individuals with elevated BP receiving antihypertensive treatment but without a formal diagnosis. These states may reflect under-recognition, incomplete diagnostic coding, or treatment for related conditions without explicit hypertension labeling (DTB 011, 012, 021, 022).

**Group 3 (G3)**: Diagnosed but untreated hypertension (DTB 100, 101, 102, 200, 201, 202). This group includes individuals with a recorded diagnosis but no pharmacological treatment and reflects potential gaps in initiation of treatment or adherence to guidelines.

**Group 4 (G4)**: Treated and controlled hypertension, including two subgroups.

**G4-1** Individuals on antihypertensive treatment despite the absence of a recorded diagnosis of hypertension, with BP in the normal range (DTB 010, 020).

**G4-2** Individuals recognized and receiving antihypertensive therapy with BP in the normal range, representing effective disease management (DTB 110, 120, 210, 220).

**Group 5 (G5)**: Treated but uncontrolled or severe hypertension (DTB 111, 112, 121, 122, 211, 212, 221, 222). This group includes individuals known to have persistently elevated BP despite treatment and/or advanced disease as well as those with resistant hypertension or a suboptimal treatment strategy.

### OUR RESEARCH

This seven-group classification captures the essential stages of care for hypertension while maintaining interpretability for clinicians, researchers, and policymakers. Importantly, these states encode not only the presence of hypertension but also the alignment, or misalignment, between clinical recognition, therapy, and biological control [2].

We investigated the clinical features of 5,129,584 patients aged 40–74 years who underwent health checkups between April 2018 and March 2019 (**Table 1**) according to these seven groups using data from the National Database of Health Insurance Claims and Specific Health Checkups of Japan [7,8]. In this preliminary study, suspected diagnoses have been excluded.

**Table 1.**
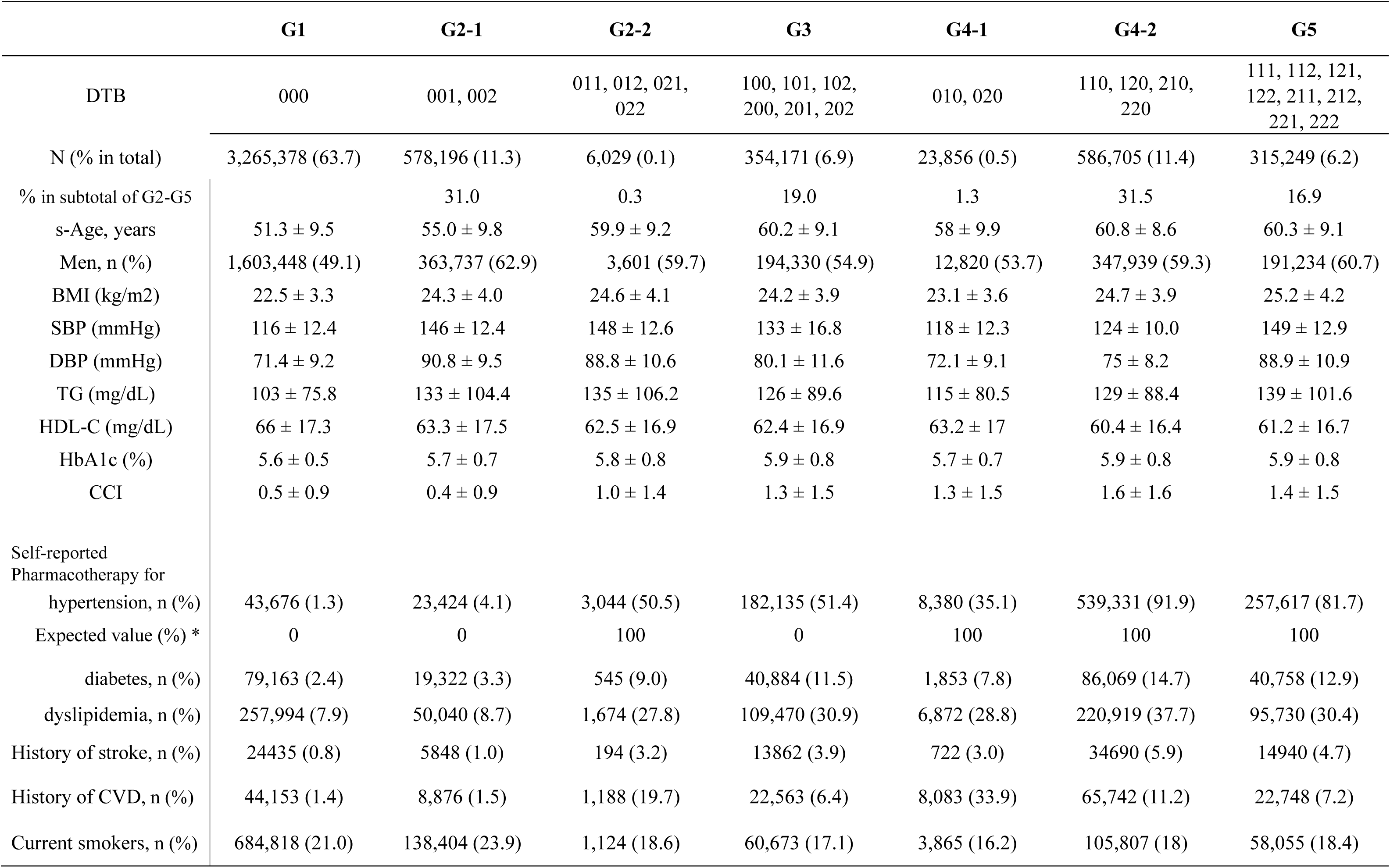
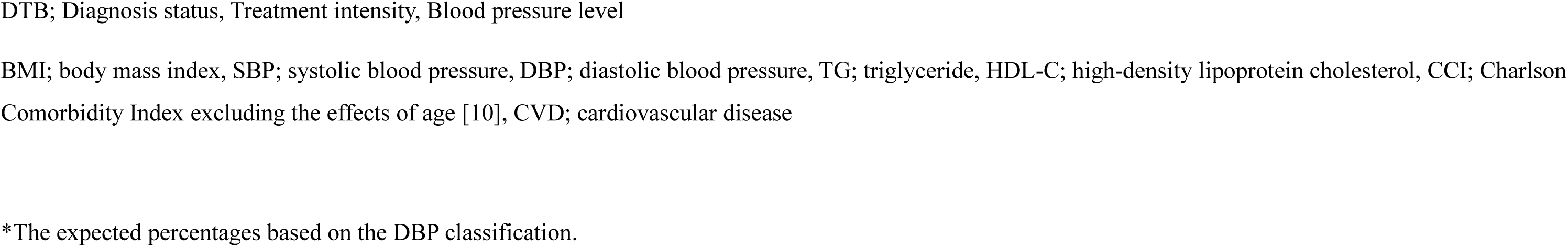
Clinical characteristics according to seven groups.

|  | G1 | G2-1 | G2-2 | G3 | G4-1 | G4-2 | G5 |
| --- | --- | --- | --- | --- | --- | --- | --- |
| DTB | 000 | 001, 002 | 011, 012, 021,<br>022 | 100, 101, 102,<br>200, 201, 202 | 010, 020 | 110, 120, 210,<br>220 | 111, 112, 121,<br>122, 211, 212,<br>221, 222 |
| N (% in total) | 3,265,378 (63.7) | 578,196 (11.3) | 6,029 (0.1) | 354,171 (6.9) | 23,856 (0.5) | 586,705 (11.4) | 315,249 (6.2) |
| % in subtotal of G2-G5 |  | 31.0 | 0.3 | 19.0 | 1.3 | 31.5 | 16.9 |
| s-Age, years | 51.3 ± 9.5 | 55.0 ± 9.8 | 59.9 ± 9.2 | 60.2 ± 9.1 | 58 ± 9.9 | 60.8 ± 8.6 | 60.3 ± 9.1 |
| Men, n (%) | 1,603,448 (49.1) | 363,737 (62.9) | 3,601 (59.7) | 194,330 (54.9) | 12,820 (53.7) | 347,939 (59.3) | 191,234 (60.7) |
| BMI (kg/m<sup>2</sup>) | 22.5 ± 3.3 | 24.3 ± 4.0 | 24.6 ± 4.1 | 24.2 ± 3.9 | 23.1 ± 3.6 | 24.7 ± 3.9 | 25.2 ± 4.2 |
| SBP (mmHg) | 116 ± 12.4 | 146 ± 12.4 | 148 ± 12.6 | 133 ± 16.8 | 118 ± 12.3 | 124 ± 10.0 | 149 ± 12.9 |
| DBP (mmHg) | 71.4 ± 9.2 | 90.8 ± 9.5 | 88.8 ± 10.6 | 80.1 ± 11.6 | 72.1 ± 9.1 | 75 ± 8.2 | 88.9 ± 10.9 |
| TG (mg/dL) | 103 ± 75.8 | 133 ± 104.4 | 135 ± 106.2 | 126 ± 89.6 | 115 ± 80.5 | 129 ± 88.4 | 139 ± 101.6 |
| HDL-C (mg/dL) | 66 ± 17.3 | 63.3 ± 17.5 | 62.5 ± 16.9 | 62.4 ± 16.9 | 63.2 ± 17 | 60.4 ± 16.4 | 61.2 ± 16.7 |
| HbA1c (%) | 5.6 ± 0.5 | 5.7 ± 0.7 | 5.8 ± 0.8 | 5.9 ± 0.8 | 5.7 ± 0.7 | 5.9 ± 0.8 | 5.9 ± 0.8 |
| CCI | 0.5 ± 0.9 | 0.4 ± 0.9 | 1.0 ± 1.4 | 1.3 ± 1.5 | 1.3 ± 1.5 | 1.6 ± 1.6 | 1.4 ± 1.5 |
| Self-reported<br>Pharmacotherapy for |  |  |  |  |  |  |  |
| hypertension, n (%) | 43,676 (1.3) | 23,424 (4.1) | 3,044 (50.5) | 182,135 (51.4) | 8,380 (35.1) | 539,331 (91.9) | 257,617 (81.7) |
| Expected value (%) * | 0 | 0 | 100 | 0 | 100 | 100 | 100 |
| diabetes, n (%) | 79,163 (2.4) | 19,322 (3.3) | 545 (9.0) | 40,884 (11.5) | 1,853 (7.8) | 86,069 (14.7) | 40,758 (12.9) |
| dyslipidemia, n (%) | 257,994 (7.9) | 50,040 (8.7) | 1,674 (27.8) | 109,470 (30.9) | 6,872 (28.8) | 220,919 (37.7) | 95,730 (30.4) |
| History of stroke, n (%) | 24435 (0.8) | 5848 (1.0) | 194 (3.2) | 13862 (3.9) | 722 (3.0) | 34690 (5.9) | 14940 (4.7) |
| History of CVD, n (%) | 44,153 (1.4) | 8,876 (1.5) | 1,188 (19.7) | 22,563 (6.4) | 8,083 (33.9) | 65,742 (11.2) | 22,748 (7.2) |
| Current smokers, n (%) | 684,818 (21.0) | 138,404 (23.9) | 1,124 (18.6) | 60,673 (17.1) | 3,865 (16.2) | 105,807 (18) | 58,055 (18.4) |

DTB; Diagnosis status, Treatment intensity, Blood pressure level
BMI; body mass index, SBP; systolic blood pressure, DBP; diastolic blood pressure, TG; triglyceride, HDL-C; high-density lipoprotein cholesterol, CCI; Charlson Comorbidity Index excluding the effects of age [10], CVD; cardiovascular disease
\*The expected percentages based on the DBP classification.

In this cross-sectional research, 64% of participants were classified as normotensive (G1), which is consistent with previous finding [2]. Hypertension-related states (G2∼G5) were observed in the remaining population (36%), close to the evidence in 2019 reported by the WHO [9], including 11% with unrecognized and untreated hypertension (G2-1), 7% with diagnosed but untreated hypertension (G3), 11% with recognized and controlled hypertension (G4-2), and 6% with treated but uncontrolled or severe hypertension (G5). Among patients with hypertension-related states (probable hypertensive patients), prevalence of untreated hypertensive patients (G2-1 and G3) was 50%, which is almost consistent with international evidence of 58% from WHO [10].

Participants who were treated in the absence of a diagnosis of hypertension (G2-2 and G4-1) were minority populations (0.1% and 0.5%). These last groups of patients had a high rate of known cardiovascular disease (33.9%) and were likely receiving antihypertensive medication for reasons other than hypertension (such as heart failure, arrhythmia, or edema) [11], which may be consistent with a lower than expected prevalence of self-reported pharmacotherapy for hypertension (**Table 1**).

The prevalence of G2-1 (11.3%) is consistent with international evidence suggesting that approximately 10–15% of adults have hypertension that remains both undiagnosed and untreated [2]. Prevalences of **G3**, **G4-2**, and **G5** in the hypertension-related states (G2∼G5) may be also consistent with Japanese evidence [12], although G3 is higher and G5 is lower than the results of the study, probably due to the bias for health checkup participation, and thereby severely hypertensive patients were not undergoing health checkups.

As a data-driven, objective analysis, we also performed a hierarchical cluster analysis using Ward’s method to classify subjects considering the standardized means of factors in **Table 1** (age, sex, body mass index, sex, serum triglyceride, high-density lipoprotein cholesterol, HbA1c, pharmacotherapy for diabetes and dyslipidemia, history of stroke and cardiovascular disease, and smoking habit) using software of SAS version 9.4. **Table 2** shows the results for the seven classifications by cluster analysis and a comparison of these classifications (**Table 1**). Clinically interpretable seven-group classification showed substantial agreement with the data-driven clusters: the simple kappa coefficient was 0.63 and the weighted kappa coefficient was 0.87 for the order of the seven groups as presented. Particularly, the major clinically relevant phenotypes, including normotensive (G1, 100 % in C1), undiagnosed hypertensive (G2-1, 100 % in C2), and successfully treated groups (G4-1, 89 % in C4), were reproduced almost perfectly by cluster analysis.

**Table 2.**
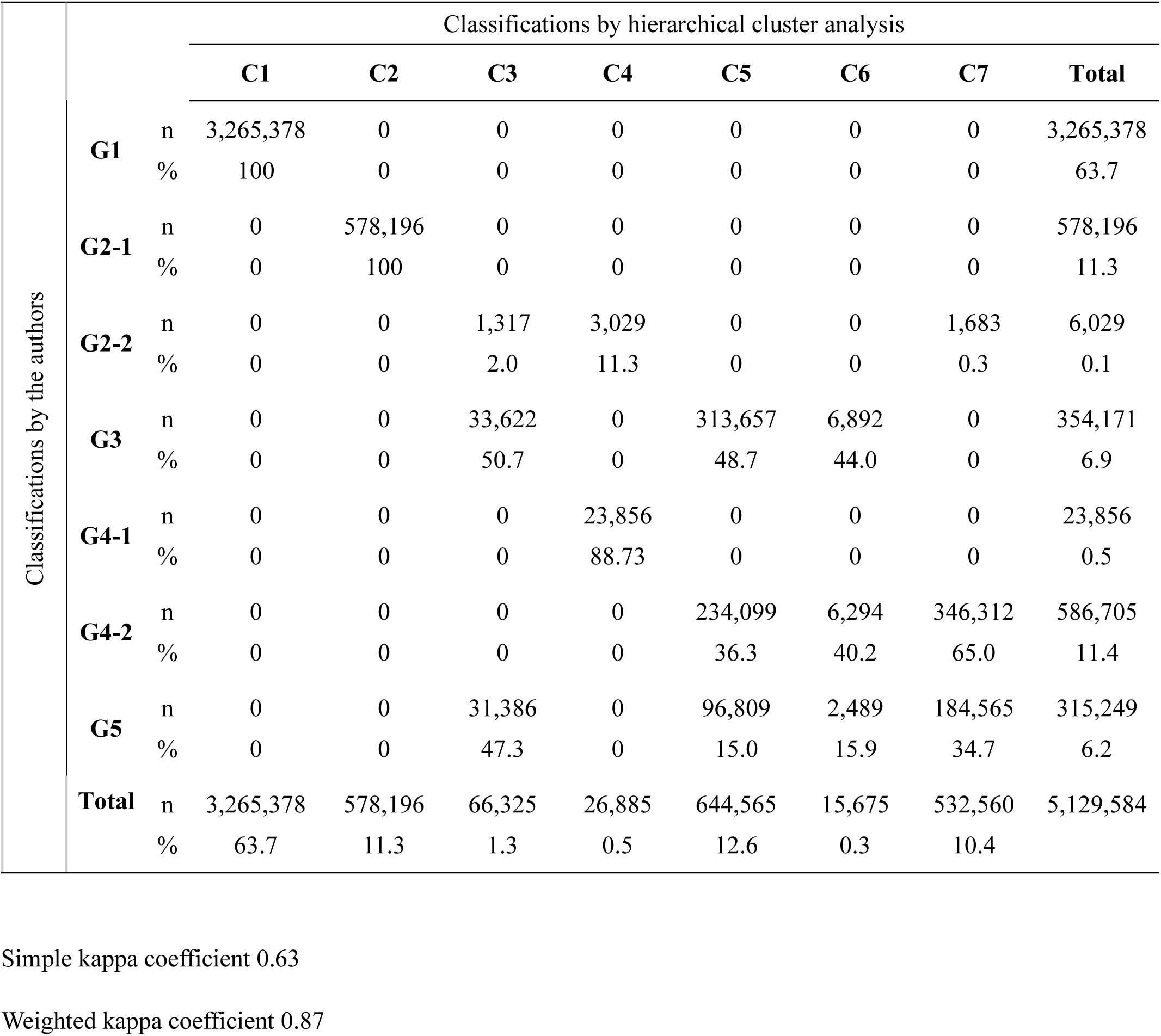
Relationship between groups classified by two methods.

Unlike G1, G2-1, and G4-1, which corresponded closely to distinct clusters, G3, G4-2, and G5 were distributed across multiple clusters. The hierarchical clustering analysis therefore identified substantial heterogeneity within these groups, suggesting that they comprise several underlying sub-phenotypes rather than single homogeneous categories. This heterogeneity may reflect differences in disease severity, patient characteristics, treatment response, and progression along the continuum of hypertension recognition, treatment, and control.

#### Sub-analysis (Sensitivity analysis)

To evaluate the influence of cardiovascular diseases other than hypertension, we repeated the analysis after excluding participants with diagnoses of angina, myocardial infarction, heart failure, arrhythmia, proteinuria, tremor, edema, and pulmonary hypertension (ICD-10, I200, I201, I208, I209, I238, I252, I210, I211, I212, I213, I219, I220, I221, I228, I229, I230, I231, I232, I233, I234, I235, I236, I241, I252, F453, G908, H350, I129, I428, I470, I480, I481, I482, I489, I490, I494, I495, I498, I499, I709, E059, I099, I110, I500, I501, I509, N391, N392, N069, N391, G250, F444, G251, G252, I500, E43, E871, I890, I270, I272). The overall distribution of the seven groups remained largely unchanged (**Table 3**). Although the proportions of G4-1 and G2-2 decreased markedly (61% and 39%), reflecting the exclusion of patients receiving antihypertensive agents for non-hypertensive indications, the prevalences of the major hypertension phenotypes (G1, G2-1, G3, G4-2, and G5) changed only modestly. These findings support the robustness of the proposed classification.

**Table 3.** Prevalence of groups after exclusion of patients with cardiovascular disease diagnoses other than hypertension.

|  | **Total** | **G1** | **G2-1** | **G2-2** | **G3** | **G4-1** | **G4-2** | **G5** |
| --- | --- | --- | --- | --- | --- | --- | --- | --- |
| N | 4,711,028 | 3,168,647 | 562,073 | 3,660 | 286,291 | 9,372 | 432,148 | 248,837 |
| (% in total) |  | (67.3) | (11.9) | (0.1) | (6.1) | (0.2) | (9.2) | (5.3) |
| % in subtotal of G2-G5 |  |  | 36.4 | 0.2 | 18.6 | 0.6 | 28.0 | 16.1 |
| Percentage decreases (%)* | 8.2 | 3.0 | 2.8 | **39.3** | 19.2 | **60.7** | 26.3 | 21.1 |

Values express n (%).
Percentage decreases from the original sample size (n = 5,129,584) due to exclusion of patients with cardiovascular disease diagnoses other than hypertension (angina, myocardial infarction, heart failure, arrhythmia, proteinuria).

#### Validity of the diagnosis of hypertension

The reliability of medical claim diagnoses is generally not high [13]. Therefore, to evaluate the validity of hypertension diagnosis in the claims database, we compared recorded diagnoses with antihypertensive medication use (**Table 4**). When the administration of one or more antihypertensive agents was used as the reference criterion, the diagnosis of hypertension showed a sensitivity of 96.5%, a specificity of 91.8%, a positive predictive value of 72.2%, and a negative predictive value of 99.2%. In addition, agreement between hypertension diagnosis and antihypertensive medication use was substantial, with a kappa coefficient of 0.78, supporting the validity of hypertension diagnosis in the claims database.

**Table 4.** Relationship between antihypertensive agent administration and hypertension diagnosis.

|  |  | No antihypertensive agent | Administration of one or more antihypertensive agents | Total n (%) |
| --- | --- | --- | --- | --- |
| No hypertension diagnosis | N | 3,852,119 | 32,200 | 3,884,319 |
|  | Row % | 99.2 | 0.8 | 75.7 |
|  | Column % | 91.8 | 3.5 |  |
| Diagnosed with hypertension, n | N | 345,626 | 899,639 | 1,245,265 |
|  | Row % | 27.8 | 72.2 | 24.3 |
|  | Column % | 8.2 | 96.5 |  |
| Total n |  | 4,197,745 | 931,839 | 5,129,584 |
| (Row %) |  | 81.8 | 18.2 | 100 |

Sensitivity, specificity, positive predictive value, and negative predictive value are 96.5%, 91.8%, 72.2%, and 99.2%, respectively, when the administration of antihypertensive drugs was considered as the hypertension criterion.
Kappa coefficient was 0.781 (95%CI, 0.780, 0.782).

## Discussion

Importantly, the proposed phenotype framework was supported by three independent lines of evidence. First, the clinically defined seven-group classification showed substantial agreement with a data-driven hierarchical cluster analysis (weighted κ = 0.87). Second, the overall group distribution remained essentially unchanged in sensitivity analyses excluding participants with cardiovascular diseases other than hypertension, demonstrating the robustness of the framework. Third, hypertension diagnosis in the claims database showed substantial agreement with antihypertensive medication use (κ = 0.781), indicating that the proposed phenotypes may be based on reliable diagnostic information. Together, these findings support the validity and robustness of the proposed classification for large-scale epidemiological research.

When hypertension is used as an outcome in a research or clinical setting, its degree and significance vary according to whether the purpose originates from a public health or clinical perspective.

For example, several target groups may be organized in descending order of population-level priority as follows: group 2 (unrecognized hypertension); group 3 (diagnosed but untreated hypertension); group 5 (severe or uncontrolled hypertension).

Alternatively, the groups may be organized in descending order from the standpoint of the clinicians as follows: group 5 (severe or uncontrolled hypertension); group 3 (diagnosed but untreated hypertension); group 2 (unrecognized hypertension).

Although only one example is presented for the classification of hypertension here, the classification should be revised according to the purpose of the research. Current findings suggest that hypertension should be conceptualized not only as a binary condition but as a multidimensional state reflecting varying degrees of alignment between clinical recognition, therapeutic intervention, and physiological control.

The clinical classification used in this study was set up for preliminary research purposes, whereas the framework proposed here has several important implications. First, it enables systematic identification of hidden high-risk populations, particularly individuals with unrecognized hypertension or persistent uncontrolled BP. These groups may contribute disproportionately to cardiovascular events despite being underrepresented in traditional analyses [14,15]. Second, it provides a basis for prioritizing interventions. By quantifying the prevalence and risk associated with each group, policymakers can target resources toward the gaps where the most impact can be made, such as improving screening programs or intensifying treatment in resistant cases. Third, it allows for comparative evaluation of healthcare systems. Differences in the distribution of these seven groups across regions or populations may reflect variations in access, quality of care, and adherence to guidelines.

Finally, the framework is generalizable. The same three-axis structure can be applied to other chronic conditions, such as diabetes and dyslipidemia, where discrepancies between diagnosis, treatment, and control are common.

There are also limitations to this type of research. First, the proposed classification framework relies on diagnostic codes [13,16], medication records [17], and clinical measurements, each of which may be subject to misclassification. Diagnostic codes may not always accurately reflect confirmed clinical diagnoses, and medications may be prescribed for indications other than the target disease. In addition, single-point clinical measurements may not fully represent an individual’s underlying disease status, potentially leading to classification errors. Second, the proposed groups may be influenced not only by biological disease processes but also by healthcare utilization patterns and access to medical care. Differences in health-seeking behavior, screening practices, diagnostic intensity, and treatment availability may affect the probability of being assigned to a particular group. Therefore, the observed distributions and transitions may partially reflect characteristics of the healthcare system rather than disease progression alone.

Future studies should validate this framework using large-scale real-world data, preferably based on repeated observations, examining its ability to predict cardiovascular outcomes and guide healthcare utilization. Nationwide databases combining longitudinal claims and clinical measurements would allow robust estimation of transition probabilities between states and enable causal inference approaches to evaluate the impact of intervention.

## CONCLUSION

Hypertension is not a binary condition but a dynamic state defined by the interaction between diagnosis, treatment, and physiological control. By integrating these dimensions into a unified framework, the model proposed here offers a more accurate representation of real-world care for hypertension. This approach shifts the focus from the presence of disease to alignment of care, highlighting actionable gaps in detection, treatment, and control. As such, it provides a foundation for improving population health outcomes and advancing the quality of cardiovascular care.

## Data Availability

All data produced are unavailable due to confidentiality reasons.

## Author contributions

K.N. contributed to the overall design of the research. K.N. and A.S. contributed to the interpretation of the initial results and discussion of the literature. K.N. and A.S. prepared the data and software for analysis. K.N. prepared the first draft of the manuscript, and both authors read and approved the manuscript. Both authors have read and agree with the version of the manuscript submitted for publication.

## Institutional review board statement

This research described here was conducted in accordance with the guidelines of the Declaration of Helsinki. The study was approved by the Institutional Review Board of the Ethics Committee of Japan Women’s University (number 513) and the MHLW of Japan (number 1320).

## Informed consent statement

Informed consent was not required in view of the anonymity of the data sourced from the MHLW of Japan as part of its nationwide program involving the provision of medical data to third parties. The study protocol is available online (https://www.jwu.ac.jp/unv/education-research/NationalDatabase.html).

## Acknowledgment

We thank Edanz (https://jp.edanz.com/ac) for editing a draft of this manuscript.

## Source of funding

This research was supported by the Research Institute at the Japan Women’s University (grant number 84).

## Disclosures

The authors declare no conflict of interest.

## Notes

There is no conflict of interest to disclose.

### Competing Interest Statement

The authors have declared no competing interest.

### Clinical Protocols

https://mcm-www.jwu.ac.jp/~NDB-K7Ps/blog/?page_id=2

### Author Declarations

The Institutional Review Board of the Ethics Committee of Japan Womens University (number 513).

